# Minimizing exposure to respiratory droplets, ‘jet riders’ and aerosols in air-conditioned hospital rooms by a ‘Shield-and-Sink’ strategy

**DOI:** 10.1101/2020.12.08.20233056

**Authors:** Patrick Hunziker

## Abstract

**Objectives:** In COVID-19, transfer of respiratory materials transmits disease but the interplay of droplet and aerosol physics, physiology and environment is not fully understood. To advance understanding of disease transmission mechanisms and to find novel exposure minimization strategies, we studied cough-driven material transport modes and the efficacy of control strategies.

**Design and methods:** Computer simulations and real-world experiments were used for integrating an intensive care setting, multi- physics, and physiology. Patient-focused airflow management and air purification strategies were examined computationally and validated by submicron particle exhalation imaging in volunteers.

**Results:** Respiratory materials ejected by cough exhibited four transport modes: long-distance ballistic, short- distance ballistic, “jet rider”, and aerosol modes. Interaction with air conditioning driven flow contaminated a hospital room rapidly. Different than large droplets or aerosols, “jet rider” droplets travelled with the turbulent air jet initially, but fell out at a distance, were not well eliminated by air conditioning and exposed bystanders at larger distance and longer time; their size predisposes them to preferential capture in the nasal mucosa, the primordial COVID-19 infection site. “Cough shields” captured large droplets but induced lateral dispersion of aerosols and jet riders. An air purification device alone had limited efficacy. A “*Shield and Sink*” approach combining “cough shields” with “virus sinks” miminized exposure to all secretions in modeling and real-life experiments.

**Conclusions:** “Jet riders” have characteristics of highly efficient respiratory infection vectors and may play a role in Covid- 19 transmission. Exposure to all droplet types can be minimized through an easily implemented “Shield and Sink” strategy.

**Graphical abstract:** 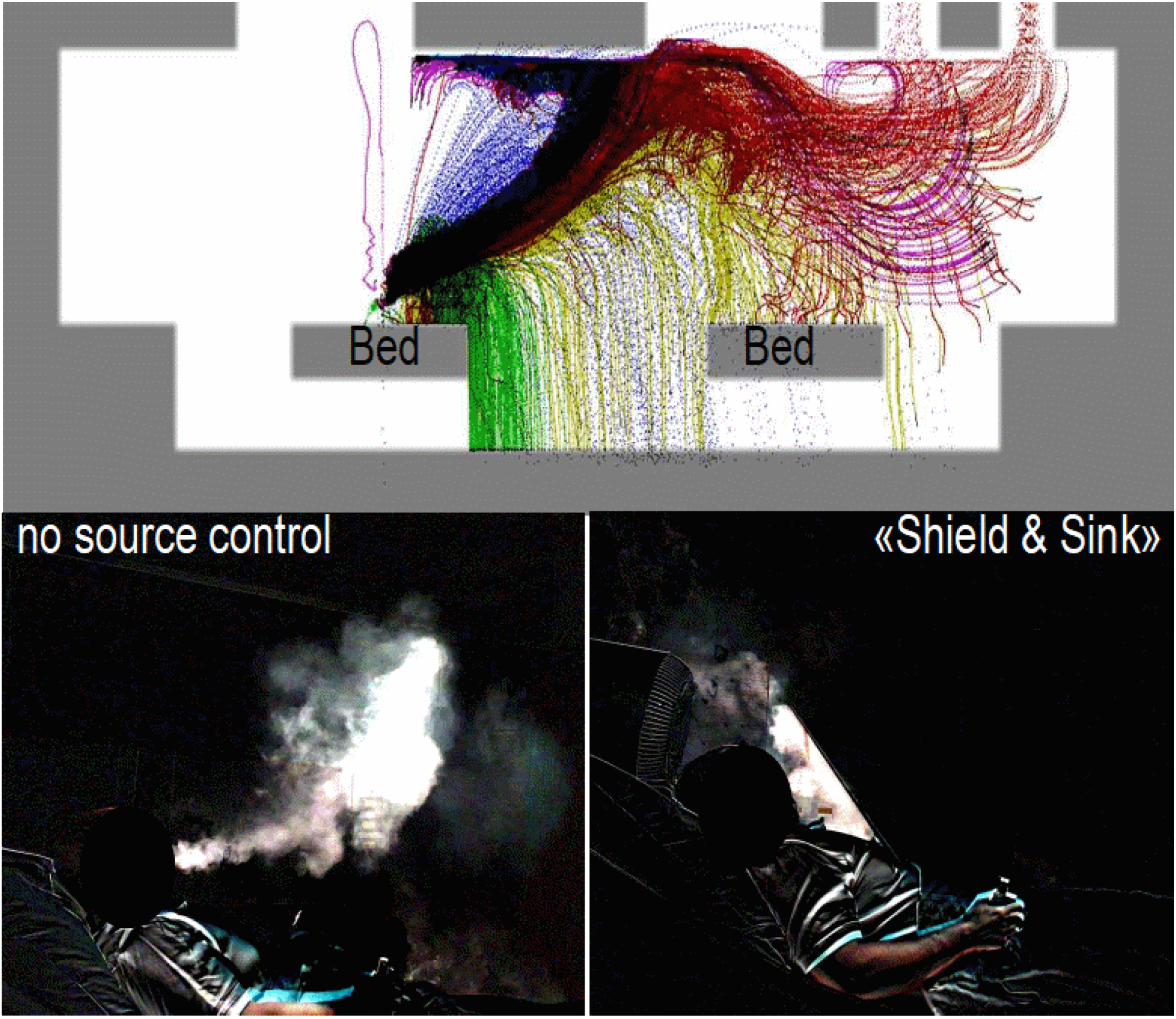

Cough-triggered dispersion of respiratory secretions of different sizes in a hospital room with air conditioning, and effective source control against dispersion by «Shield & Sink» strategy.

**Article Summary – strengths and limitations:**

- The novel “Shield and Sink” approach for source control of droplets, “jet riders” and aerosols emitted by coughing patients permits significant reduction of the transfer of such potentially infectious materials in a hospital setting.
- As a rapidly and inexpensively implementable infrastructural measure, it has the potential of contributing to mitigation and control of the Covid-19 pandemic.
- The study is performed in a hospital setting and the Covid-19 pandemic, but the implications and principles of “Shield and Sink” air management for respiratory disease transmission control are generalizable to other settings and infectious agents.

## Introduction

### Role of droplets and aerosols in Coronavirus transmission

Droplet transmission from coughing or sneezing is considered the dominant mode in COVID-19 but current evidence also suggests a role of aerosol-mediated virus transmission^1^,^2^ rendering understanding and control of emitted respiratory materials of all sizes desirable. Arguments for a role of aerosols include transmission by asymptomatic persons^3^,^4^,^5^, epidemiologic observations in SARS-CoV-2^6^,^7^,^8^, air sample analysis^9^,^10^ in SARS- SARS-COV-2^11^ and SARS-COV-1^12^, and general observations in virus and bacterial aerosol transmission^13^,^14^,^15^. Controling aerosol transmission may be an adjunct to avoiding droplet transmission for COVID control^16^,^17^, as recently emphasized by many experts^18^.

### Expiratory droplets, aerosols and submicron particles: Physiological origin, physical aspects, room distribution and persistence

Droplets of various sizes are produced when air moves across wet respiratory surfaces during respiring, coughing, sneezing, talking or singing. Droplet number and size mix depend on activity and will influence weight and infectious agent content; also, droplet motion^19^ is size-dependent, ranging from ballistic transfer to slip flow to free molecular flow. The multiphase air jets in coughing or sneezing contain various droplet sizes; in small droplets termed aerosols, aerodynamic drag dominates gravity, such that they tend to travel with the air rather than to falling to the ground; in contrast, larger droplets move ballistically due to larger mass and stronger gravitational impact and may transition to aerosols upon water evaporation. Understanding droplet and aerosol mediated disease transmission therefore needs consideration of production, initial acceleration, droplet/aerosol characteristics and behavior of the environment like air motion.

Real-life imaging of droplets and aerosols produced by coughing and respiration is possible^20^,^21^,^22^,^23^, but methods have practical limitations. E-cigarette smoke has submicron particle size comparable to respiratory aerosol subpopulations^24^,^25^ and is accessible to imaging, so that E-cigarette smoke tracking might enhance understanding aerosol clouds and allow validating theoretical predictions.

### Source control and personal protective equipment

The current emphasis in Covid-19 transmission control is on social distancing, testing, isolation of affected individuals and their contacts, and use of personal protective equipment (PPE). PPE like masks and gowns confer significant protection, but the relevance asymptomatic and presymptomatic transmission and the phenomenon of super spreaders calls would call for broad use of PPEs, and if not worn to the full extent, may lead to partial protection only. Improved source control might contribute to transmission risk reduction particularly in settings where widespread use of full PPE is not feasible.

### Aim of the study

We aimed at in-depth understanding of transmission of potentially infectious secretions from coughing persons to their environment in a hospital context, based on cough physiology, real-life room architecture and air conditioning. Using computational and real-life experiments, we further aimed at developing effective source control measures that minimize transfer of such materials and may thus minimize indoor COVID-19 transmission risk.

## Methods

Methods include modeling of room architecture, air flow and cough physiology combined with real-world experiments in a hospital setting. Hospital rooms involved in COVID-19 patient care in the University Hospital Basel, Switzerland, were recreated as volumetric digital models, equipped with patient beds, monitors, desks, air conditioning system, cough shields and air sinks.

### Air flow modeling

A computational multi-physics model was based on room geometry and standard fluid dynamics modeling of air flow, based on the hydrodynamic principles described by Bernoulli at our university in 1738^26^, formalized as Navier-Stokes equations and implemented as an open source computer library, as described in the supplemental material. Modeling included pressure, flow, aerosol content, temperature and buoyancy. Room ventilation was parameterized according to the existing air conditioning system, with settings ranging from minimum regulatory recommendations for ICU rooms [> 40 m^3^/h/person + 100 m^3^/h/patient^27^,^28^] up to 10 room air exchanges per hour for a given room. Patient airflow was modeled as air emanating from the mouth of a patient in a semi-recumbent bed position (i.e., cough jet direction upwards with 30° forward tilt) and optional lateral head rotation. Normal respiration and biphasic flow patterns^29^ for cough were modeled, with “exhalation”, “weak cough” and “strong cough” modeled as peak airflow velocities of 1.5m/s, 10m/s and 20m/s, respectively. A “breathable zone” (typical caregiver head location), was defined from 1/3 to 2/3 of room height, with particle density in this zone as surrogate for caregiver exposure, while particles sticking to surfaces or floating near the floor or the ceiling were considered “not breathable”.

### Droplet and aerosol modeling

Modeling of droplet and aerosol distribution and dynamics was done in two complementary ways: particle modeling and diffusion modeling.

In *particle modeling*, individual droplets and aerosols were modeled as spheres moving in the air flow that was modeled by fluid dynamics. Physical particle modeling included starting point, ejection velocity at mouth level, ejection angle, aerodynamic drag exerted by the surrounding moving air, including gravitation, random effects and Brownian motion. Droplets were generated in numbers proportional to orifice velocity, in size clusters with mean diameters of 1mm, 100µm, 10µm, 1µm and 0.1µm +/- 20%, corresponding to observed droplet sizes in cough^30^ and sneezes, down the size of a single virus particle. Terminal velocity for each droplet due to gravity was determined as a function of the Reynolds number. Dynamics for each droplet within the fluid dynamics field were modeled, using both, separate runs for each size cluster as well as combined runs to understand differential distribution and fate of droplets. Droplet impact on surfaces led to sticking of the particle to said surface. Droplets sticking to a surface or exiting the room through exhaust pipes or door were counted.

In a *convection*/*diffusion model*, complementary to the particle analysis, air contamination by aerosols was modeled as continuous diffusion model, with origin of the solute at the mouth, using particle size to compute the diffusion coefficient. Modeling details are given in the supplement.

### “Virus Shield and Sink”

Virus shields consisted of bed-mountable transparent polycarbonate plates of size 45cm*35cm to 60cm*60cm, with their corresponding digital model, positioned in front of the patient’s head and angulated such that the dominant cough direction hits the shield approximately in right angle. A “Virus sink” consisted of a commercial air purifier device (Toom Model 30W, REWE Zentral AG, Köln, Germany) having an air inlet diameter of 32cm, a HEPA filter, was capable of purifying up to 350m^3^/h, and its corresponding digital model, having H13 Hepa filter characteristics, positioned at the cranial bed end, with the air inlet near the patient head.

### Real-life imaging of submicron particle distribution on coughing

Two habitual E-cigarette smokers were positioned in an ICU bed in the modeled room, instructed to inhale the smoke of their E-cigarette and then to voluntarily cough with moderate strength, and were filmed for digital submicron particle imaging without and with the “Shield and Sink” equipment in different combinations.

### Patient and public involvement

The study was triggered by discussions between physicians and ICU nurses on strategies to minimize in- hospital virus transmission during the ongoing Covid-19 pandemic.

### Ethics

The responsible Ethics Commission Northwest and Central Switzerland EKNZ (Req-2020-01362) ruled that the study is a not clinical study and does not require Ethical Committee permission. The persons visualized in figures and supplements gave written informed consent for participation and consented to their images and videos being published for scientific purposes.

### Dissemination to participants and data sharing

Data have been made available to participants; data will be available as supplementary files.

## Results

### Modeling: Room geometry and ventilation (Figure 1)

Room geometry, room installation and air flow due to the air conditioning system (A/C) using standard A/C settings are shown in figure 1. A downdraft near the patient’s leg location from the A/C inflow to the outlet vents near the door drives a horizontal air motion component even in stationary flow conditions. Flow was almost stationary (compatible with the Reynolds number ∼2000) in standard A/C settings but changed to an oscillating pattern with increased turbulence (Reynolds number ∼5000) at 10 air exchanges per hour.

**Figure 1:**
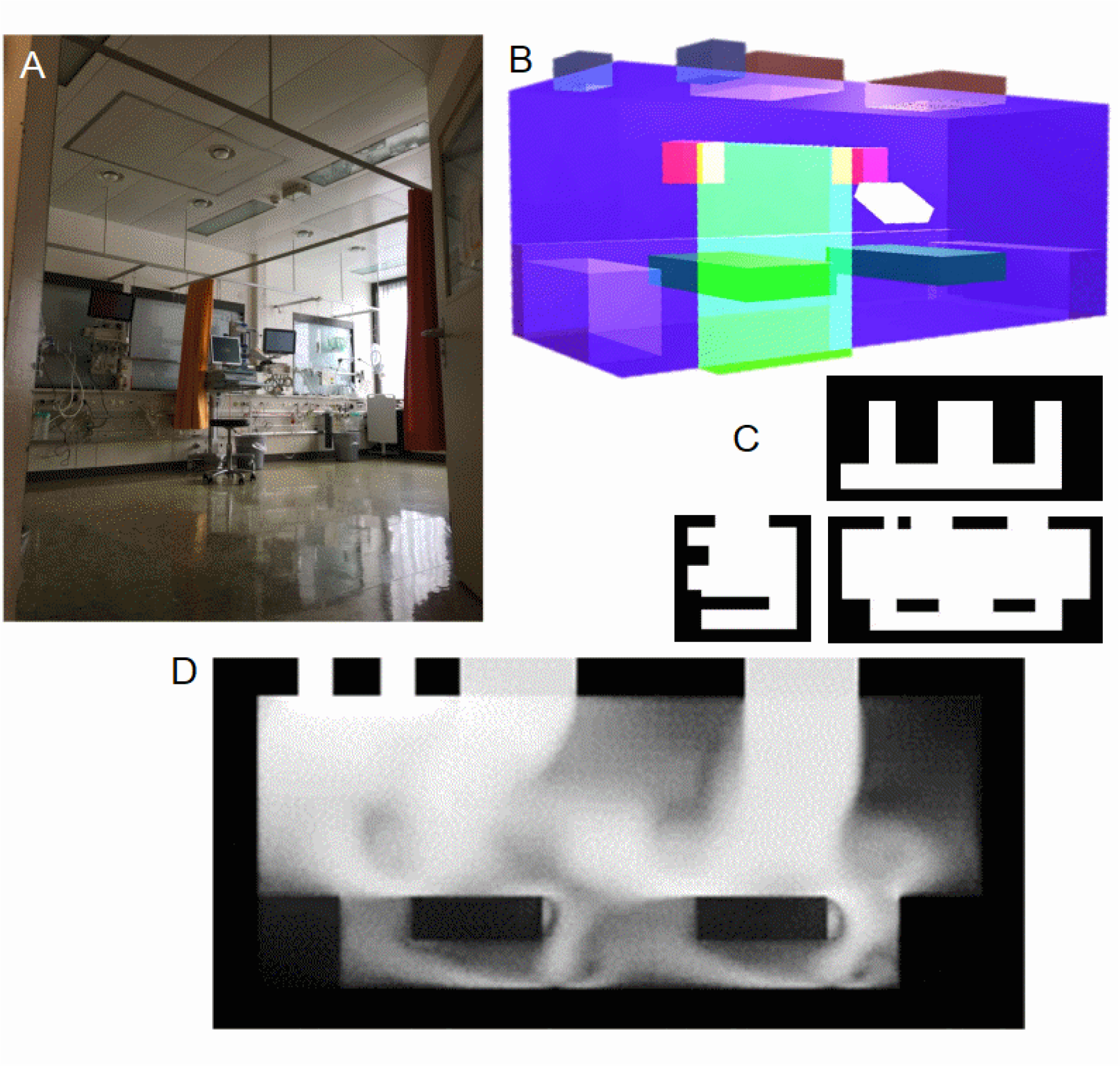
Room geometry and air flow. A) ICU room. B) 3D model of ICU room: Dark green: beds; rose: monitors; brown: air conditioning inlets; dark gray: air conditioning outlets; bright green: door; white: cough screen mounted at patient bed: transparent blue: nurse work benches. C) Room cross sections: Top, front and side view. D) Example of modelled air flow pattern induced by air conditioning at 4 room air exchanges per hour. Brightness encodes sum of velocities in side projection. Note the 2 inflows (top middle, top right), the 2 outflows (top left) and the turbulent flow patterns around the beds (bottom half) with air flow, e g., from the right bed to the left bed and the nurse workbench (right border)

### Distribution of ejected droplets and aerosols in modeling (Figure 2)

Four characteristic dynamic behaviors were documented: In the first seconds of a cough, ballistic ejection of 1mm droplets up to 4 and 6 m distance during moderate and strong coughs was observed in the particle model, capable to reach the contralateral wall, nurse workplaces and adjacent beds, depending on head rotation. Notably, the upward/forward direction of the cough jet due to the semi-recumbent position essentially doubled the reach of ballistic droplets compared to a horizontal jet direction. Droplets in the 0.1mm size range started ballistically but were quickly slowed down by aerodynamic drag and, attracted by gravity, fell down within 1 m around the patient head, such that after the first 3 seconds, most droplets from 1 to 0.1mm had hit a surface and were no longer in the air. Droplets in the 10µm size range behaved differently: due to the small size and a high drag/gravity force ratio, they travelled with the air jet for several meters before falling out due to gravity from the slowed down air convection; distance traveled by these droplets even surpassed most large ballistic droplets; they were the main population sprayed on a neighbor bed in a specific setting. A fourth population, the droplets of size 1µm down to 0.1µm showed classical aerosol behavior and travelled throughout with the air convection, not falling out at the studied range of ventilation settings. Such aerosols followed complex trajectories within the room determined by A/C induced air flow patterns, and depending on cough direction and ventilation settings, reached most room locations within a few minutes, before being removed by exiting through the ventilation exhaust, sticking to a wall, or, with the door opened, by leaving through the door to central ICU workplaces.

**Figure 2.**
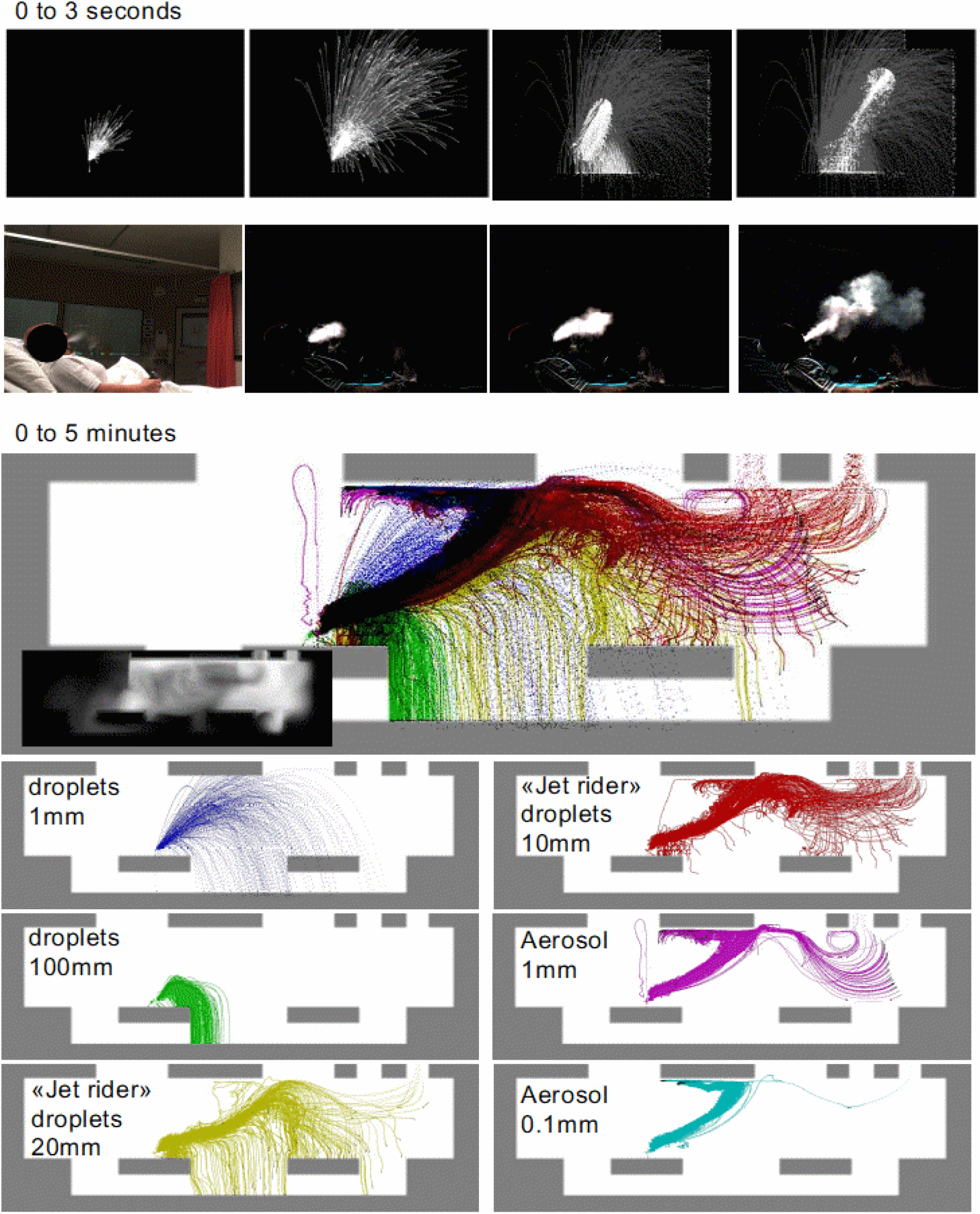
Distribution of ejected droplets and aerosols after a cough. Top: Ejection of droplets in first 3 seconds (row 1: modeled, row 2: in particle imaging) after a cough. A-D) modeled, including droplets from 1 mm down to 0.1 μm. White dots indicated current droplet position, gray traces indicate trajectory of droplet since start. E-H) particle imaging using E-cigarette smoke (submicrometer particles) and digital subtraction imaging (F-H). Bottom: Transport of various sized droplets within 5 minutes after a cough.

The convection-diffusion model yielded complementary results consistent with the aerosol particle modeling and showed that within minutes, in most locations of the room (except downstream of the air conditioning inlet), concentration of aerosols build up and are only slowly cleared (10-20min) by air conditioning, implying that a higher cough frequency leads to accumulation of aerosols.

### Exposition to various droplet fractions at “breathable locations” over time (Figure 3)

The time course of particles in the “breathable zone” of the room is shown in figure 3. Larger droplets were quickly sprayed across space but hat all but disappeared from the air at 3 seconds, either sticking at walls, floor or beds. Aerosols in the 0.1 to 1 micrometer range persisted for 20 minutes in significant proportion in the breathable zone and showed recirculation, but due to their propensity to rise towards the ceiling through the buoyancy of the warm exhaled air (where the air conditioning exhaust is located), exposition in the breathable zone was lower than for “jet riders” (droplets in the 5-40 micrometer size range) that dominated the breathable zone air up to 300 seconds and resulted in large exposition-time integrals, a surrogate for exposure intensity.

**Figure 3:**
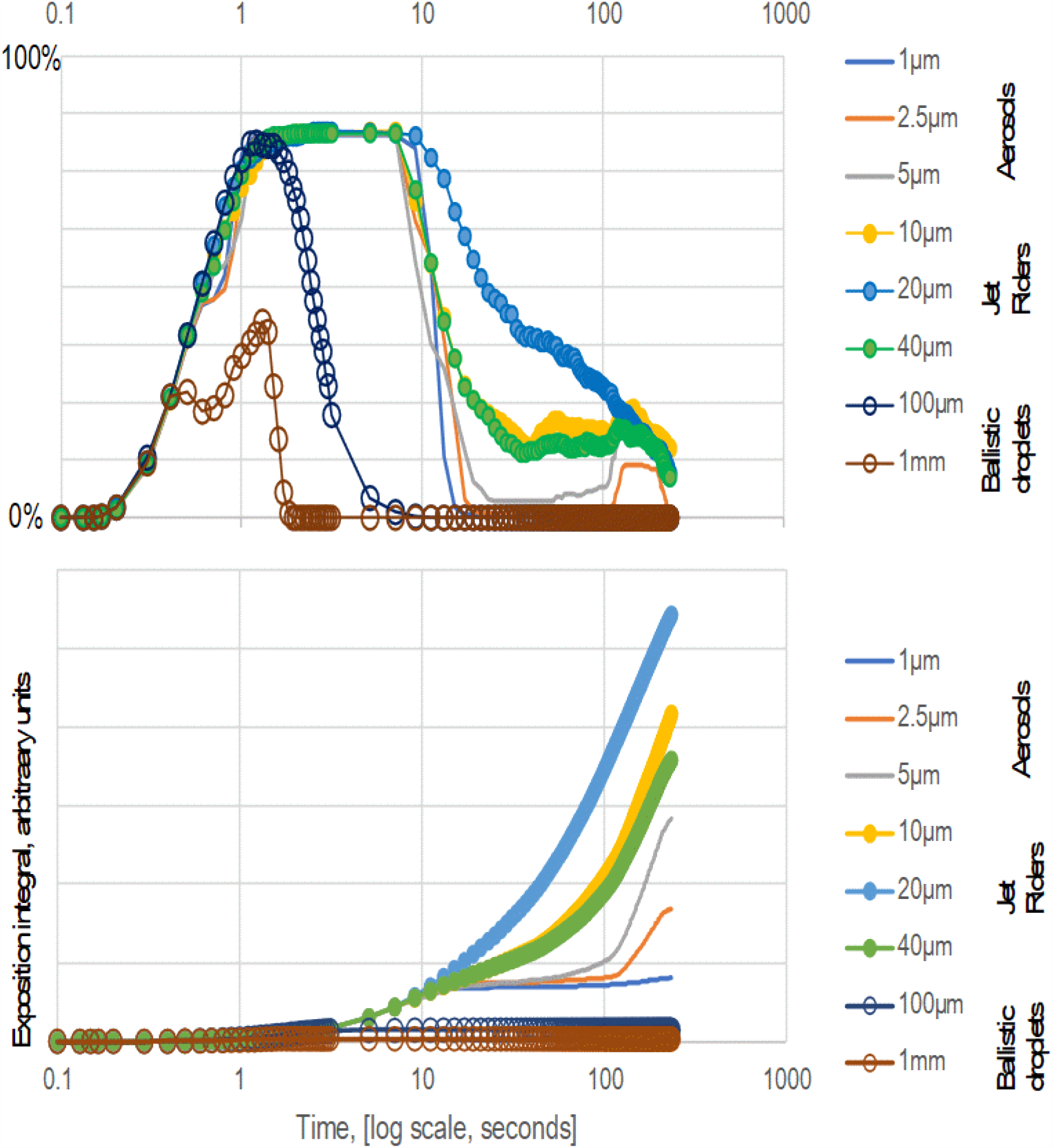
Time-course of droplets and aerosols in breathable space after a cough. Time course of droplets/particles as a function of their size, measured as their proportion present in «breathable space», i.e. in the air at a room elevation of. Ballistic droplets (0.1-1mm) spray the environment of a source but are gon within a few seconds.

### Droplet and aerosol capture strategies in model: (Figure 4)

*Virus shields* positioned in front of the patient could captured all large droplets with trajectories suited to reach distant locations. However, air jet splitting and redirection was observed with such shield, leading to lateral aerosol plumes moving in direction of nurse workplaces and the adjacent patient bed.

**Figure 4:**
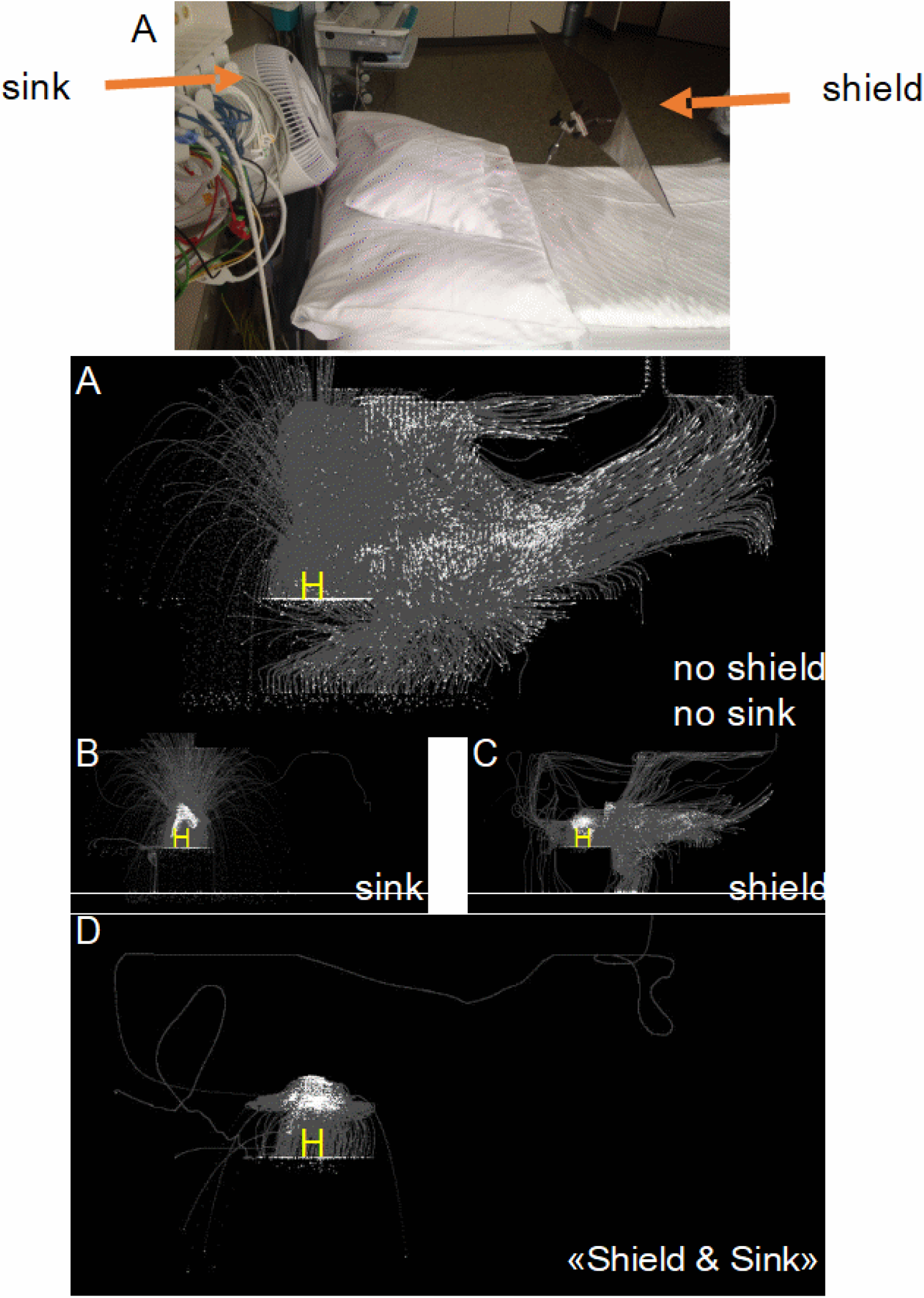
Source control strategies to avoid dispersion of droplets and aerosols. Efficacy of source control strategies for cough-related particle dispersion, modelled. Top: “Shield and sink” installed in an ICU bed Bottom: Trajectories (gray) and locations at 5 minutes (white) of droplets and aerosols emitted by a single cough. H indicates the patient head position, i.e. the jet origin. A) unprotected cough, rapidly spraying the environment with ballistic droplet and dispersing jet riders and aerosols across the room. B): Installing an air purifying «sink» captures aerosols but is incapable to stop ballistic droplet spray C) shielding the patient’s face stops ballistic droplets but disperses jet riders and droplets in multiple directions. D) Only a «Shield and Sink» strategy is capable to quantiatively eliminate respiratory secretions from being transferred to the environment: almost all material sticks to the shield or is absorbed by the air purifying sink

A *virus sink*/air filtration device positioned near the patient’s head and driven at maximum performance was neither capable to capture the larger droplets nor the majority of the aerosol produced, because the jet emanating from the mouth was sufficiently fast and strong to overcome the small flow velocity field produced by the device.

In the novel *Shield & Sink* strategy, a shield and a sink were combined based on parameters identified in modeling. Modeling showed that at sufficient sink air flows, the shield bends the streamlines of the cough air flow such to keep the aerosol plume in proximity to the sink device, so that it can be quantitatively captured and filtered by the device. Stepping up from a H13 to a higher performant H14 Hepa filter had a negligible additional effect on the results, but higher cough jet velocities increased the proportion of “escaping” aerosol while higher air sink velocity decreased it (data not shown). The added value of air disinfection strategies, e.g. by ultraviolet light was not examined but the results suggest that its potential for incremental benefit to “Shield & Sink” is small.

### Droplet and aerosol capture strategies in real-life scenario (Figure 5)

Assessing the behavior of exhaled sub-micrometer particles by digital submicron particle imaging of ejected E-cigarette smoke confirmed computational predictions: Coughing rapidly produced a large particle plume, discernibly by eye up to 2 meters in the first seconds, followed by slow further movement. Installing a shield in front of the face led to jet splitting and redirection. Using an air filtration device near the patient’s head was incapable to quantitatively remove the aerosol. However, in the “Shield and Sink” strategy, the shield slowed the aerosol jet such that the plume was aspirated and filtered to a visually full degree by the sink device. The video clips in the supporting material show the performance of droplet and aerosol capture strategies in modelled and real-life scenarios.

**Figure 5:**
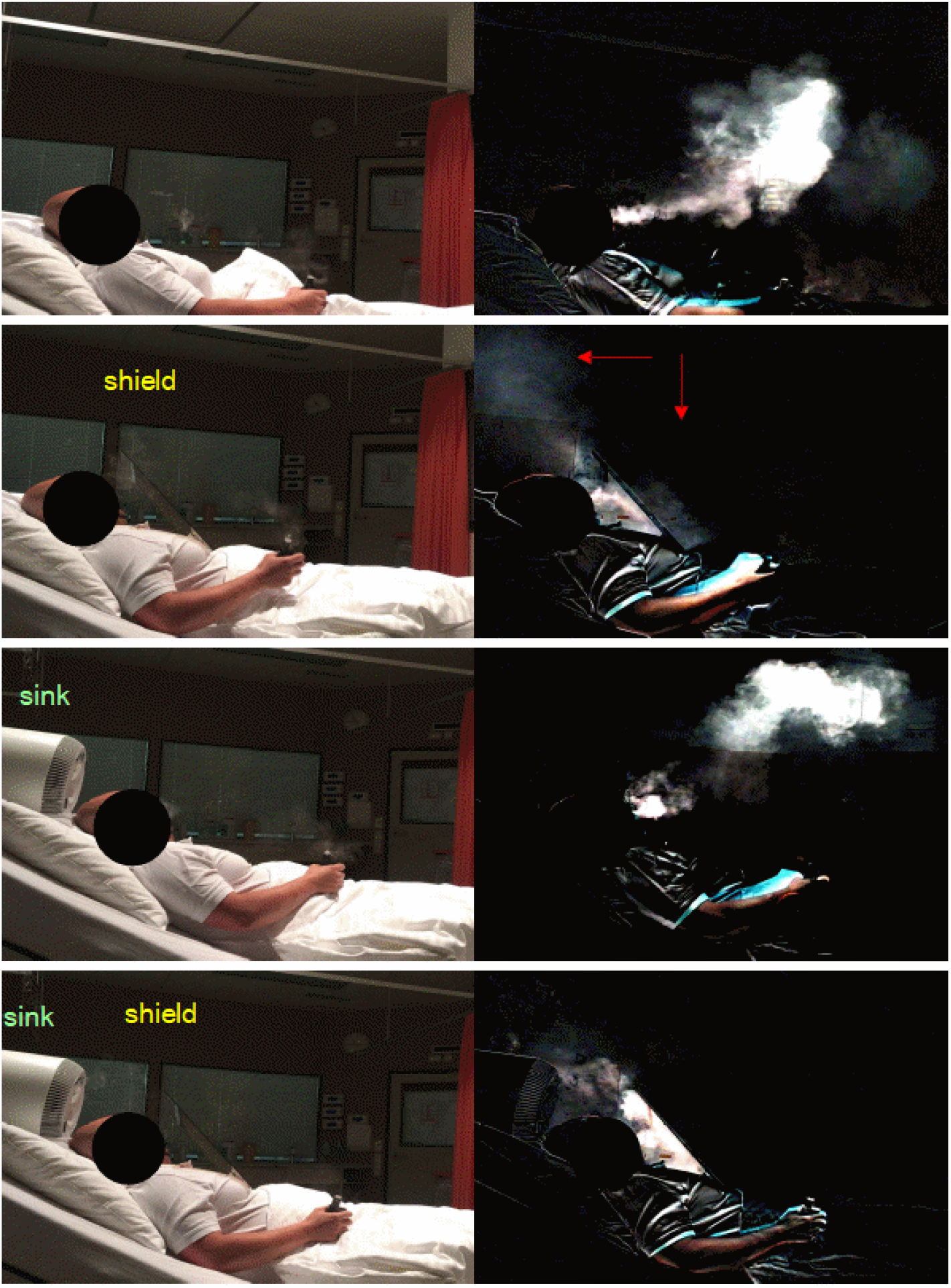
Real-life particle imaging of droplet and aerosol capture strategies. Efficacy of source control strategies for cough-related particles in particle imaging. Top row: Ejected small particle cloud within 3 seconds after a cough. Particles are from E-cigarette smoke that is dominated by submicrometer particles. 2nd row: A «cough shield» in front of the patient’s face leads to dispersion of the particle jet in multiple directions. C) An air purification device in proximity to the patient head is incapable to capture the particles, becauase the cough jet velocity is higher than the device-induced air flow. D) «Shield and Sink» strategy with a shield in front of patient head and an air sink (a commercial air purification device with a Hepa filter) near the patient head. Note the near quantiative removal of cough related particles.

## Discussion

In the context of the ongoing COVID-19 pandemic, this study analyses the spread of potentially infectious materials produced by respiratory events like coughing and examines the value of source-control measures for avoiding droplet- and aerosol transmission of respiratory materials to bystanders during hospital care of patients. It quantitatively analyzes distribution, recirculation and out-of-room transfer of droplets and aerosol produced by coughing and breathing through computational modeling of patient activity, room architecture and air conditioning and validates the findings by exhaled particle cloud imaging. Extending prior knowledge, it describes four different and potentially medically relevant modes of droplet transport: 1) dominant ballistic (long distance fallout) 2) ballistic-drag (short distance fallout), 3) floating-gravity dominated (longest distance fallout), and 4) fully floating (no fallout).

The novel “Shield and Sink” approach introduced here is a patient-centered strategy to transmission avoidance by creating per-patient aerodynamic settings, an approach that proves highly effective and achieves near-complete elimination of droplets and aerosol of all classes near the patient head in modeling and real-world experiments, suggesting a potential for strongly reducing exposure of care persons and nearby patients.

### The quest for source control measures to stop infection propagation

In view of the persistence^31^ of SARS-CoV2 in aerosols with an estimated half-life of 60min, personal protective equipment (PPE) for minimizing droplet and aerosol inhalation is widely used in direct COVID patient care. However, the occurrence of asymptomatic carriers and super-spreaders renders each untested patient entering a hospital a potential source of infection. Rapid testing of each patient and use of full PPE for each untested individual may not be realistic and economically feasible in all settings at a global scale, raising the question if, and to what degree, suited *source-control measures* could prevent transmission of infectious material to caregivers, other patients and other bystanders.

### Determinants of droplet and aerosol propagation

We found that the spread of potentially infectious material from the airways to the environment is highly variable and determined by patient characteristics (coughing strength, head position), droplet size, room geometry and ventilation conditions. Even in ventilation conditions fulfilling current hospital architecture norms, droplet spread reached neighbor patients and nurse workplaces; prolonged aerosol circulation was observed, but increasing ventilation settings was found to potentially increasing room air turbulence, i.e. undesired air mixing.

For droplets in the 1 millimeter range, we observed ballistic transfer up to 5-6 meters in strong and 4 m in moderate strength cough jets. The longer reach of droplets compared to the historical 1-2 m rule for droplet transmission has several reasons: First, in ICU and hospital room patients, the frequently used semi- recumbent position^32^ with head elevation ≥30° directs the cough jet obliquely upwards and thereby maximizes a ballistic trajectory; also, such droplets initially travel with the air jet (i.e., less drag), benefit from upward buoyancy of the warm expired air, and dry up slower due to the expired air humidity, enabling them to reach farther than an isolated water droplet horizontally emitted into non-moving dry air. Droplets in the 0.1 mm diameter range also behaved ballistically, but their smaller kinetic energy led to quick slowdown by drag as soon as they left the cough air jet, and being pulled down by gravity, their reach was limited to approximately 1m around the head. Typical aerosol behavior (traveling with the surrounding air, flowing around obstacles) was observed for droplets starting at the 1 µm down to the 0.1 µm ranges, consistent with literature; the negligible impact of gravity on such very small droplet combined with the higher temperature of the exhaled air explained their tendency to rise towards the ceiling and being eliminated through the air conditioning exhaust. “Jet rider” droplets, smaller than ballistic droplets but larger than classical aerosols, appeared to have particular importance:

### “Jet Riders” with distant fallout

Notably, the size fraction of droplets in the 10-20 (or 5-40) µm range displayed a behavior that does neither fit in the “ballistic” nor in the “aerosol-type” patterns: While it travels in the early, fast, warm, turbulent and buoyant air jet like an aerosol, the gravitational pull becomes dominant in the slower moving air with less turbulence, leading to gravitational fallout at a distance, thereby spraying objects like a neighbor patient bed or nurse workplaces, reaching on average even further than the large ballistic droplets, a finding that may be highly relevant because smaller droplets carry a significant amount of infectiosity^33^,^34^. Unfortunately, this is also the particle size range where the nose, the primordial location of SARS-CoV-2 infection, catches particles highly effectively^35^.

While classical fluid dynamics, aerodynamics and droplet physics are well established scientific fields, it appears thus that in the complexities encountered in the interaction of human pathophysiology, viral spread, droplet physics and room air conditioning, some important and interesting phenomena are not yet fully explored, appear relevant for human health and thus merit further in-depth study that may contribute to solving urgent societal problems.

### Hospital air conditioning

Air flow induced by A/C was an important driver of aerosol transmission to adjacent patients and nurse working locations and recirculation of contaminated air. Doors also played a role: typically, only a small minority of hospital rooms is equipped for negative pressure settings but default ventilation in our institution in patient rooms, builds up a slight positive pressure; when patient room doors remain open or are opened as needed for patient care, transfer of aerosols to the central nurse workplace occurred (data not shown). Increasing air flow rate from rates on one side led to more aerosols leaving the room per time interval, but on the other side induced more turbulent room air flow at the given geometry, favoring air mixing and thus potentially transporting aerosol-loaded air from non-breathing zones towards breathing zones; therefore, the benefit of just increasing ventilation thus has limitations that depend on the specific characteristics of a room and A/C system.

### Cough shields

Cough “shields” were partly effective in stopping forward-directed cough droplets but led to a redirection of the air stream carrying aerosols, including in the direction of adjacent beds and nurse workplaces at the side of the patient bed, an effect which is not desired. Also, air flow around obstacles is a well-known physical phenomenon^36^ that limits the effectiveness of such shields for aerosol plumes.

### Air filtering

Air purification by filtering with a high flow, HEPA filter equipped commercially available air filtering device, was examined. We found in modeling as well as in real-world experiments (E-cigarette smoke distribution) that such an air filtering device alone, even when positioned near the patient head, only has a limited capabililty for directly and quantitatively removing droplets/aerosols from the cough jet because the jet velocities away from the device were sufficiently high to overcome the modest pressure/velocity gradient produced by the device. Non patient centered air filtering has been shown to contribute to reduction of overall aerosol load in recirculating contaminated room air^37^, but in such that setting, an aerosol concentration half-time in the range of 6-12 minutes was found, similar to the aerosol removal in our context by standard A/C. In such devices, quantitative aerosol removal is dominated by air flow, directing flow towards the filter, rather than Hepa filter class (H13: 99.95%; H14: 99.995% particle removal efficacy).

### “Virus shield and sink” – a novel, effective, per-patient airflow handling approach

To achieve near-complete elimination of droplets and aerosols of all sizes from 1mm down to 0.1micrometer from the cough jet and expired air, a novel per-patient airflow handling approach was designed, inspired by the findings above, and was validated in a real-life scenario.

Such a per-patient airflow management includes capturing large, ballistic, forward directed droplets by a shield that at the same time works as a jet redirector, limiting the forward velocity of the air jet and redirecting the aerosol plume in a way that it is amenable to being captured by a “virus sink”, a strategically placed aur filtering device behind the patient’s head driven at velocities tailored to bend the streamlines of the aerosol jet towards the device and quantitatively filter the aerosol. This setup was capable to capture cough-related respiratory materials from millimeter-sized droplets down to submicron particles in a quantitative fashion.

### E-cigarette smoke test as simple surrogate test for source control measures

The efficacy a specific “Shield and Sink” setup tailored to a specific location and filter device that may differ in geometry and air flow capacity from the one described here is a amenable to simple qualitative function testing by the E-cigarette smoke test described here, thus representing a real-world quality control and training tool.

### Limitations

Clinical proof of infection reduction by preventive measures will require large-scale clinical trials that cannot easily be blinded. In the ongoing COVID pandemic, time available for such trials is limited, and clinical trial evidence for most prevention activities will remain sparse. As the approach proposed here is non-invasive, is associated with limited expense, and the current study focuses on a plausible surrogate marker, namely spread of various-sized materials produced by coughing, such a strategy may be clinically introduced with little delay, yielding a safe time window to further in-depth study of the still controversial role of aerosols in COVID transmission by additional methods, e.g. aerosol nucleic acid testing and cell culture infectiosity of exhaled materials.

### Importance

The novel “Virus Shield and Sink” approach for source control of droplets, “jet riders” and aerosols emitted by coughing patients permits significant reduction of the transfer of such potentially infectious materials in a hospital setting. As a rapidly and inexpensively implementable infrastructural measure, it has the potential of contributing to mitigation and control of the COVID pandemic.

## Supporting information

Supplement Modelled-No-Source-Control

Supplement Modelled-Source-Control

## Data Availability

The data are available on request from the author for scientific purposes with suited attribution and as long as the manuscript is not published, an agreement on confidentiality.

## Contributorship

All parts of the work performed by P.H.

## Supplemental material

### Architectural Modeling

Architecture modeling was done within a multiphysics environment (“A2” Framework, “Matrix” library developed in a joint project of the University of Basel and ETH Zurich) by creating a 3D vector geometry model using the software module “VolumeRaytracer”). Regular multiresolution finite difference grids from size 512×256×256 down to 32×16×16 were then overlaid on the volume data for modeling.

### Fluid dynamics modeling

The fluid dynamics model was based on the same multiphysics framework, module “VolumeNavierStokes” and was typically run on the 256×128×128 grid, with a 16-fold oversampling for particle displays. Fluid dynamics used a Backward Time Centered Space approach and modeled fluid advection, diffusion and divergence using a standard operator splitting approach under the assumption of noncompressibility, also incorporating solute injection at the mouth orifice, solute diffusion as well as heat transfer (orifice exit temperature 37°C) and buoyancy. Aerosol production (modeled in this part as solute injection) was proportional to orifice exit velocity. Diffusion coefficients for spheres of 10um, 1um and 0.1um were 3.0E-8, 3.0E-7 and 3.0E-6, respectively. Adaptive time stepping was used to limit fluid advection within a step to at most one grid element, resulting in time steps down to 2ms. Measurement probe volumes could be placed within the model to monitor the changes of a local parameter over time. An overall Reynolds number as predictor of turbulent flow conditions was estimated using the room extents and the overall flow, although precise, local Reynolds number in such complicated geometries cannot easily be determined.

### Particle modeling

Particle modeling was done using the same framework, with an object-oriented modeling of each particle in a manner allowing overlay of the particles on the fluid dynamics framework. Specifically, individual droplets and aerosols were considered as spheres with the density of water moving in the air flow that was modeled by fluid dynamics. Physical effects in sphere modeling included ejection velocity at mouth air speed, aerodynamic drag exerted by the surrounding air, gravitational force, and Brownian motion, each including some random variation. Droplets originated at random positions within the orifice, were generated in numbers proportional to the orifice velocity, in size clusters with mean diameters of 1mm, 100µm, 10µm, 1µm and 0.1µm +/- 20%, corresponding to observed droplet sizes in cough and sneezes down the size of a single virus particle. Initial velocity corresponded to the fluid dynamics velocity field with added Gaussian noise (sigma=25%) to velocity vector components. Drag and terminal velocity for each droplet was determined based on the drag equation, switching to the terminal velocity creeping flow solution^38^ when Reynolds number was <1. Dynamics for each droplet were modeled, in separate runs for each size cluster and in combined runs to understand differential distribution and fate of droplets. Droplet impact on surfaces led to sticking of the particle to said surface. Droplets sticking to a surface or exiting the room through exhaust pipes or door were counted as inhalable in the exposure statistics.

### Cough physiology modeling

The airflow model was similar to the one described in PLOS. In short, on coughing an air jet exited from an orifice (area 5cm^2^), with a temporal cough flow profile modeled as two-phase event with a first, stronger but short (peak at 0.25 sec) air emission, overlapping with a somewhat longer (peak at 0.5 sec) but less strong air emission, whereby each component was modeled as a cropped (zero to pi) sinus wave, parameterized such that for “strong cough”, “moderate cough” and “normal exhalation” mouth orifice peak velocities approximated 20m/s, 10m/s, and 2.5m/s, respectively. Video clips were produced that show the time course of material dispersion after a cough: in modelled video clips the first three seconds in first 3 seconds in realtime, followed by or a 5-minute time window by in time-lapse. In real-life video clips, the first few seconds after a cough are shown.

### Submicron particle exhalation imaging

Submicron particle exhalation imaging was achieved by first filming a particle exhaust plume originating from exhaled E-cigarette smoke with a stationary iPhone 6s camera under strong back-lighting illumination conditions in a darkened room, followed by digital subtraction image processing whereby the initial frame is subtracted digitally from subsequent frames (custom software setup, including ImageJ software^39^).

---

**Supplemental Figure:**
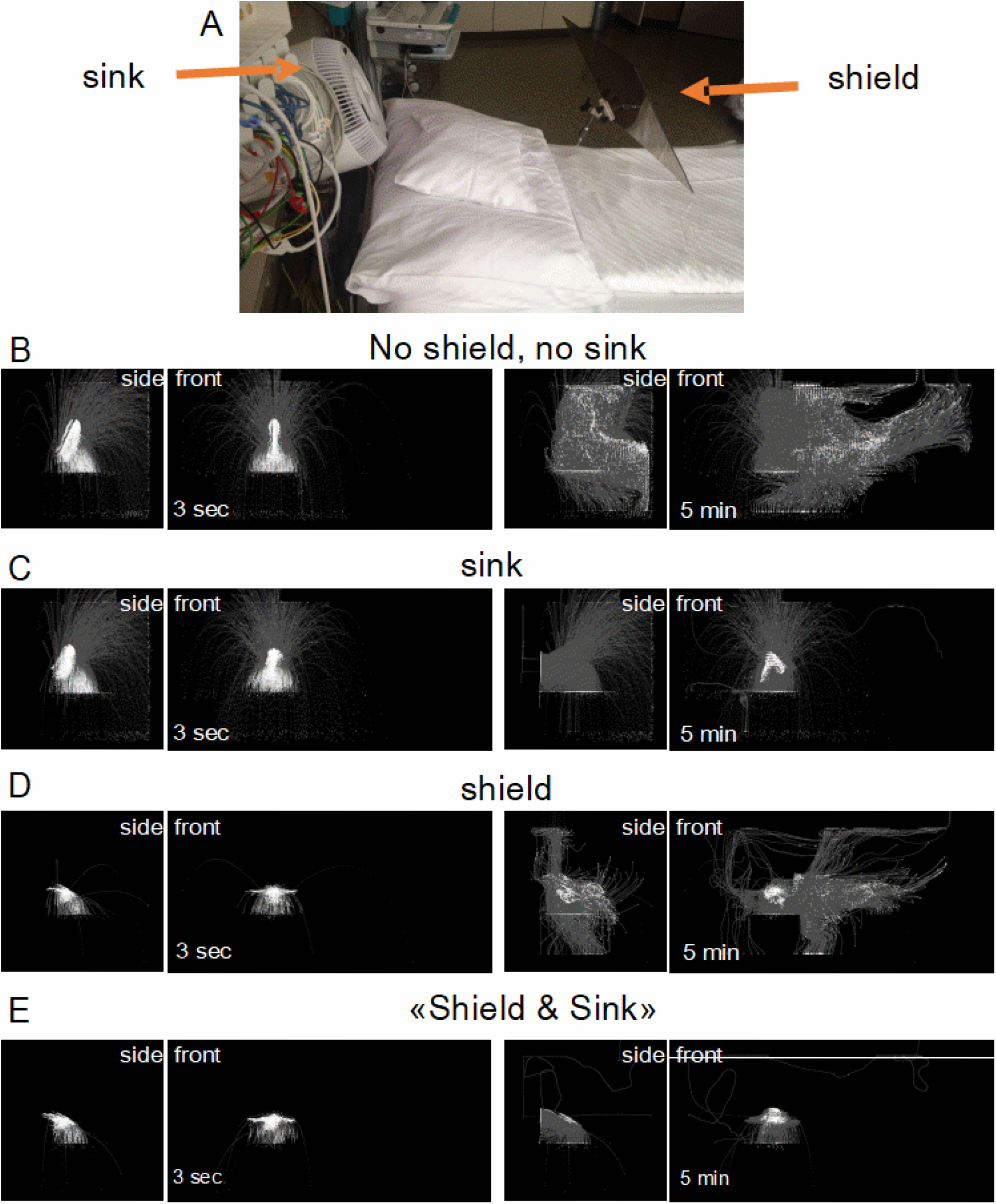
“shield and sink1”- differential effects on early droplet spray and subsequent aerosol distribution. A) An air sink and a cough shield mounted on an ICU patient bed. B) unprotected cough. rapidly spraying the environment with ballistic droplet and dispersing jet riders ana aerosols across the room. C): Installing an air purifying «Sink» captures aerosols but is incapable to stop ballistic droplet spray. D) shielding the patent’s face stops ballistic droplets but disperses jet riders and droplets in multiple directions. E) Only a «Shield and Sink» strategy is able to quantiatively eliminate respiratory secretions from being transferred to the environment.

